# Health and economic impact of geographically prioritized long-acting PrEP delivery in southern and eastern Africa

**DOI:** 10.64898/2026.02.24.26345396

**Authors:** Adam Akullian, Jeffrey W. Imai-Eaton, Monisha Sharma, Hasina Subedar, Michelle O’Brien, Geoffrey P. Garnett

## Abstract

**Background:** Long-acting injectable HIV pre-exposure prophylaxis (PrEP), including Lenacapavir, has the potential to accelerate HIV incidence declines in eastern and southern Africa (ESA). However, high product and delivery costs and constrained budgets necessitate efficient prioritization strategies to maximize impact and achieve cost-effectiveness.

**Methods:** We used district-level HIV incidence estimates published by UNAIDS to estimate the direct health and economic impact of prioritizing Lenacapavir delivery according to geography, age, and sex across 837 districts in 11 high-burden ESA countries. Infections and disability-adjusted life years (DALY) averted, number needed to treat (NNT), cost per DALY averted, and price thresholds to achieve cost-effectiveness were estimated across geographic prioritization scenarios. Cost-effectiveness was assessed against a $500 per DALY averted threshold, assuming $5,000 discounted lifetime HIV treatment costs and 10 DALYs per HIV infection. Sensitivity analyses varied Lenacapavir costs (commodities + delivery) per person per year (pppy) ($125 versus $55), DALYs per HIV infection (7.5), and the risk differentiation among those who uptake long-acting PrEP.

**Results:** HIV incidence varied substantially across ESA, with 50% of new infections in districts containing less than 20% of at-risk adults. Lenacapavir cost-effectiveness varied accordingly, with high-incidence districts exhibiting substantially lower NNT and higher price thresholds for cost-effective delivery. In high-incidence districts, [>5/1,000 person-years (py)], of South Africa, Mozambique, Lesotho, and eSwatini, Lenacapavir would be cost-effective at $50-100 pppy. In South Africa, at annual cost $55 pppy, Lenacapavir was cost-effective in all 52 districts when provided to women aged 15–24 years with incidence exceeding twice the district average and could reach approximately 18–20% of new infections while covering 4% of the full HIV-negative adult population aged 15–49 years. Geographically optimized prioritization in South Africa with minimal age and risk-group stratification achieved efficiency comparable to country-level prioritization to high-risk groups and key populations (∼20% incidence reduction with 3–5% coverage). Impact and cost-effectiveness were sensitive to assumptions about risk heterogeneity.

**Conclusions:** Lenacapavir impact and cost-effectiveness varies substantially across geographic settings, driven primarily by variation in HIV incidence. Simple incidence-based models can identify where universal provision to certain demographic groups is both impactful and cost-effective, particularly in high-incidence districts and age groups.

## Introduction

Long-acting pre-exposure prophylaxis (LA-PrEP), including the 6-monthly injectable Lenacapavir, provides an opportunity to accelerate declines in new HIV infections in Eastern and Southern Africa (ESA)^1–3^. However, with increasingly constrained and less ring-fenced budgets for HIV treatment and prevention, implementation strategies must achieve meaningful impact at costs that represent good value relative to other opportunities to invest health resources. In launching and scaling new prevention tools, there is often an inherent trade-off between affordability and impact. Relatively high-cost HIV prevention, including Lenacapavir, is often only determined to be cost-effective when used by those with the highest risk of acquisition, and available budgets may constrain the scale at which implementation is affordable to the health system. Yet, even as the total number of new infections is much lower than in previous decades, most new infections in generalized epidemics still occur among a large population with limited distinguishing risk factors^4,5^. This raises questions about where, how, and to whom the newly approved Lenacapavir should be delivered.

Several validated HIV risk prediction scores have been previously developed to identify those at highest HIV risk based on individual-level demographic and behavioral co-variates^6–10^. These tools, however, have demonstrated mixed performance^8^, often missing many of those who subsequently acquire HIV infection, particularly young women^9^. Therefore, the utility of individual risk differentiation tools in prioritizing PrEP provision may be limited. Furthermore, implementing a risk prioritization approach adds personnel time, costs, and complexity to product delivery^11^. Risk screening may also stigmatize populations identified as “high-risk”^12^, potentially leading to lower uptake among those who would benefit most, further reducing the sensitivity of these tools in reaching persons most likely to acquire HIV.

Simpler PrEP delivery strategies, such as a universal offer of PrEP in high-incidence settings, have been proposed as an alternative to risk-prioritization models^13,14^. Universally offering PrEP overcomes the limitations of risk-screening and allows the user to decide whether and when to take PrEP based on their own risk perception, which can increase coverage among those at highest risk and maximize impact of PrEP. There is potential for this approach to be both efficient and impactful. Those who self-assess elevated risk tend to have higher PrEP initiation compared to those identified via a risk scoring algorithm^15^, and self-assessed risk may closely align with an individual’s empirical risk^16^, resulting in efficient impact on reducing incidence in real-world settings^13^. Re-analysis of data from an observational study embedded within the ECHO trial^13,17^ shows that when oral PrEP was offered to all enrolled women, the HIV risk of those who took up oral PrEP was approximately six times higher than those who did not (corresponding to a >2-fold higher risk in oral PrEP users compared to the overall population). Offering PrEP to broad demographic groups without restriction in high-incidence geographies may streamline HIV prevention without losing efficiency.

Most mathematical models that have evaluated the health impact and cost-effectiveness of Lenacapavir in ESA have utilized complex dynamic transmission models^4,18,19^. Such models are resource-intensive to run and rely on complex data and assumptions for parameterization. Simpler tools may provide similar policy conclusions while requiring fewer parameters and being user friendly and transparent for decision makers^20^. Here, we use estimates of incidence at the district level across ESA in a simple model to estimate the direct health impact and cost-effectiveness of Lenacapavir provision. We develop a heuristic approach for Lenacapavir prioritization using a minimal set of demographic, epidemiological, and costing parameters to estimate cost-effectiveness and impact of various geographic prioritization scenarios. Our results can inform epidemiological thresholds where Lenacapavir could be cost-effectively offered universally versus risk-prioritized.

## Methods

We expressed the direct health impact and cost-effectiveness of long-acting PrEP using simple mathematical formulas, based on a previously described framework^20^. These equations were used to estimate direct infections averted using inputs on HIV incidence in the absence of PrEP, PrEP efficacy, duration of protection, and population coverage. Second, costs and health impacts were assessed based on the annual per-person cost of Lenacapavir and estimates on the average avoided lifetime healthcare costs and DALYs lost per HIV infection (See appendix for equation inputs).

Estimates for HIV prevalence, incidence, and population at risk by district, age, and sex were produced using the Naomi model^21^, and reported by countries to UNAIDS as part of 2025 UNAIDS annual epidemiologic estimates^22,23^. The geographic scope included 837 districts in 11 ESA countries. Naomi synthesizes data on HIV prevalence and ART coverage from population-based surveys and routine HIV service delivery to estimate district-level HIV prevalence, ART coverage, and HIV incidence. District variation in HIV incidence is modelled proportional to population prevalence of viraemia, and sex and age patterns are based on incidence rate ratios from national epidemic estimates using the Spectrum model^24^.

Model parameters and values are shown in Table 1. Our base-case calculations assumed that persons using Lenacapavir have the average age- and sex-specific HIV incidence rate for their district in the absence of PrEP. We also considered a scenario that assumes universal PrEP offer would lead to uptake among those who have more than twice the average HIV risk in their district based on results from a PrEP implementation study embedded within the ECHO trial^13^. This assumption is consistent with findings on incidence among participants in clinical HIV prevention trials, which is ∼2–3 times higher than average, potentially indicative of higher risk individuals self-selecting to participate into HIV prevention programs^25^. To model individual-level variation in HIV risk, we parameterized risk heterogeneity (*r*) assuming a continuous gamma distribution: *r* ∼ *Gamma*(*shape k, scale θ*), where *k* is fixed and represents within-district skew, and scale θ is calculated such that the distribution mean reproduces age and sex stratified district-level incidence. Gamma distributions have been used to represent skewness in the distribution of transmissions and acquisitions in a population for several infectious diseases^26^. In HIV epidemic models, Gamma distributions may approximate the skewness in the number of sexual partners per person per year across a population^27^, where a small number of individuals have high activity versus a larger number with average or low-to-moderate activity. For our analysis, we assumed risk follows an exponential distribution with a shape parameter of 1 to represent a moderately skewed distribution of risk, following previous work^28^. In the case of a Gamma with shape 1, more than 50% of infections fall within the top 25% of the population with the highest risk and those in this population have, on average, twice the risk of the general population (Figure S1). We estimate only the direct heath impact of Lenacapavir in preventing HIV among those taking it and do not account for secondary transmissions averted.

**Table 1.**
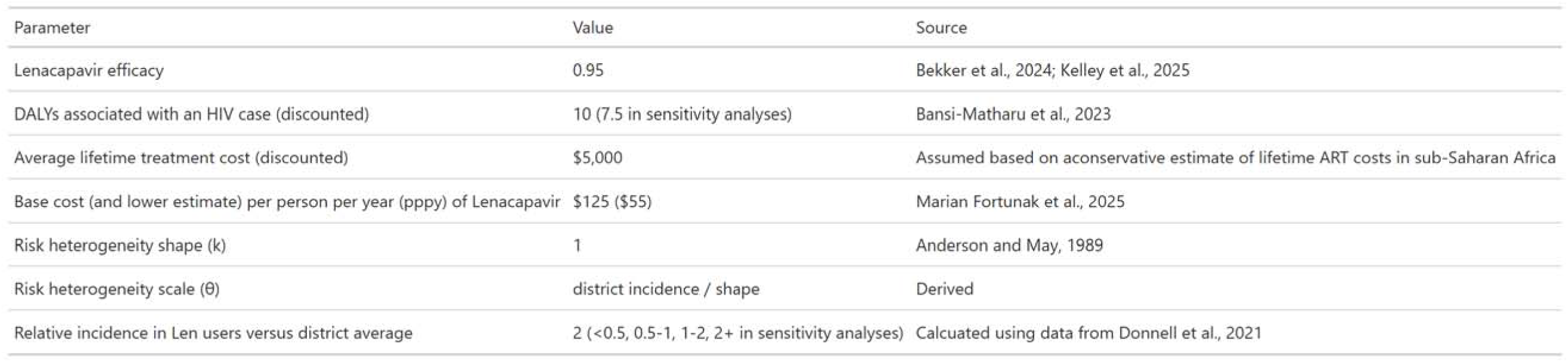
Model parameters and values.

| Parameter | Value | Source |
| --- | --- | --- |
| Lenacapavir efficacy | 0.95 | Bekker et al., 2024; Kelley et al., 2025 |
| DALYs associated with an HIV case (discounted) | 10 (7.5 in sensitivity analyses) | Bansi-Matharu et al., 2023 |
| Average lifetime treatment cost (discounted) | \$5,000 | Assumed based on a conservative estimate of lifetime ART costs in sub-Saharan Africa |
| Base cost (and lower estimate) per person per year (pppy) of Lenacapavir | \$125 (\$55) | Marian Fortunak et al., 2025 |
| Risk heterogeneity shape (k) | 1 | Anderson and May, 1989 |
| Risk heterogeneity scale ($\theta$) | district incidence / shape | Derived |
| Relative incidence in Len users versus district average | 2 (<0.5, 0.5-1, 1-2, 2+ in sensitivity analyses) | Calculated using data from Donnell et al., 2021 |

We assumed Lenacapavir efficacy of 95% for 6 months based on phase III efficacy trials, which found 100% protection against HIV acquisition among cis-gender females and 96% efficacy in sexual minority men and women^29,30^. In our baseline analyses, we assumed a Lenacapavir cost of $125 per person per year (pppy), which reflects the fully loaded costs of Lenacapavir provision including drugs, HIV testing, personnel, and consumables, oral loading dose, and demand generation. We also modeled a lower cost scenario at $55 pppy, representing a fully loaded cost of commodity and delivery at a reduced commodity price of $40 pppy^31^. Scenarios were determined to be cost-effective if the cost per DALY averted was <$500 per DALY averted^32^ and cost-saving if the costs of providing Lenacapavir were less than the discounted treatment costs averted.

To quantify the cost and health losses avoided per HIV infection averted, we assumed a discounted $5,000 lifetime treatment cost per person, reflecting a conservative estimate of the marginal cost of long-term ART provision in high-coverage settings^33^. We assumed an average of 10 DALYs associated with HIV, reflecting the years of life lived with HIV and the years lost due to premature mortality, based on similar ranges from HIV costing models, derived from dividing total DALYs averted by infections averted^34^. Alternate costing and DALY parameters were used in sensitivity analyses to estimate a lower bound cost threshold for various delivery strategies as described below.

### Sensitivity analyses

We conducted sensitivity analyses to assess the impact of assumptions regarding DALYs associated with an HIV infection, lifetime treatment costs of a PLHIV, and the risk segment of the population most likely to uptake PrEP based on a risk distribution curve. We ran additional models under more conservative assumptions of 7.5 DALYs associated with an HIV infection (Table 1). For the risk-heterogeneity assumption, we tested the sensitivity of our results across four groups represented as quantiles of the risk distribution (0–25%, 25–50%, 50–75%, and 75–100%), corresponding to groups with a relative difference in risk from the district-level mean of <0.5, 0.5–1, 1–2, and >2, respectively. We assume that the upper quantile, representing 25% of the population with the highest risk (i.e., >2-fold higher incidence), will be the most likely PrEP adopters.

## Results

HIV incidence rates in 2024 varied substantially across the 837 districts in 11 eastern and southern African (ESA) countries (Figure 1). High-incidence districts (≥5 infections per 1,000 person-years among adults aged 15–49 years) were concentrated primarily in southern Africa, particularly in South Africa, Mozambique, Lesotho, and eSwatini (Figure 1A) and accounted for a disproportionate share of new HIV infections relative to the population at risk. Districts with incidence ≥5/1,000 person-years contributed 34.1% of new infections while containing only 11% of the adult population at risk.

**Figure 1.**
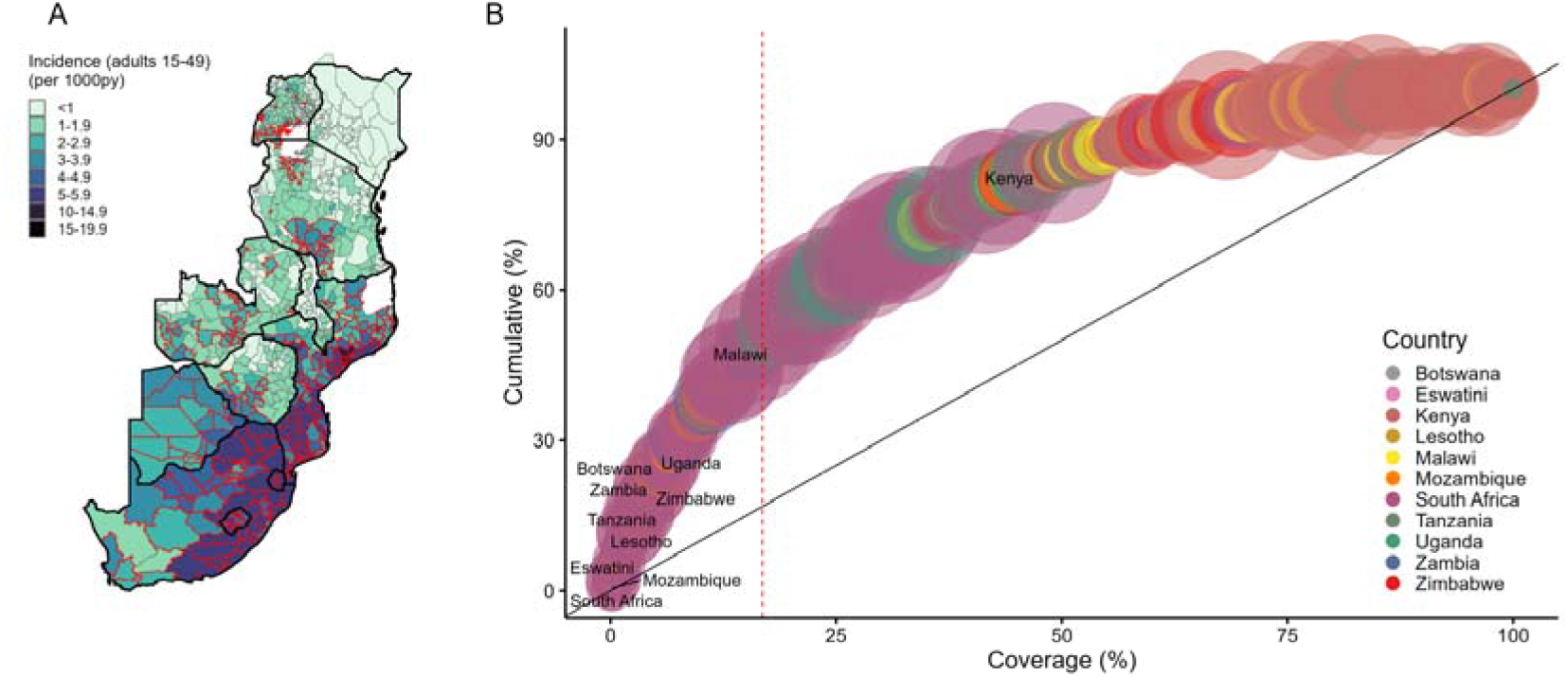
(A) District-level incidence (per 1000py) among adults aged 15–49 years, with red highlighted districts indicating where incidence of any age (15–24, 25–34, 35–49) and sex (men and women) group is above 3.8 per 1000py, representing 50% of new infections and 17% of the HIV negative population. (B) Cumulative percent of new infections with increasingly inclusive age and sex stratified HIV negative population. Points are ordered from highest to lowest incidence; color represents country and point size represents the age- and sex-specific HIV negative population size

In contrast, districts with low HIV incidence (<1/1,000 person-years) were predominantly located in eastern Africa and contributed a much smaller fraction of new infections (7.5%) relative to their population size (32.8%). This marked geographic heterogeneity translated into strong concentration of infections within a small fraction of the population (Figure 1B). Across ESA, 50% of new infections occurred in districts containing less than 20% of the population, highlighting the potential efficiency gains of geographically prioritized prevention strategies (Figure 1).

District-level HIV incidence also varied substantially by age and sex (Figure S2). Among women, incidence was typically highest in those aged 20–24 years and lowest among those aged 35 years and older, although in several high-burden districts in South Africa incidence peaked among adolescent girls aged 15–19 years. Among men, incidence was consistently low in the 15–19-year age group and higher among those aged 20 years and older.

The cost-effectiveness of Lenacapavir varied widely across districts with different HIV incidence (Figure 2A). Districts with higher HIV incidence exhibited lower numbers needed to treat (NNT), lower costs per DALY averted, and higher maximum fully loaded pppy prices at which Lenacapavir would remain cost-effective (assuming a willingness-to-pay threshold of $500 per DALY averted, $5,000 in discounted lifetime ART costs, and 10 DALYs per HIV infection). In districts in South Africa, Mozambique, Lesotho, and eSwatini where incidence ≥5/1,000 py, fewer than 200 adults 15–49 would need to receive Lenacapavir to avert one HIV infection. Under base-case assumptions, Lenacapavir would remain cost-effective at prices between $50-$100 pppy. In contrast, in lower-incidence districts, NNT values exceeded 1,000 and cost per DALY averted frequently surpassed accepted thresholds. When delivered to young women (15–24) with average district-level risk, Lenacapavir was cost-effective between $50-$100 pppy in most districts in South Africa, eSwatini, and Mozambique (Figure 2b).

**Figure 2.**
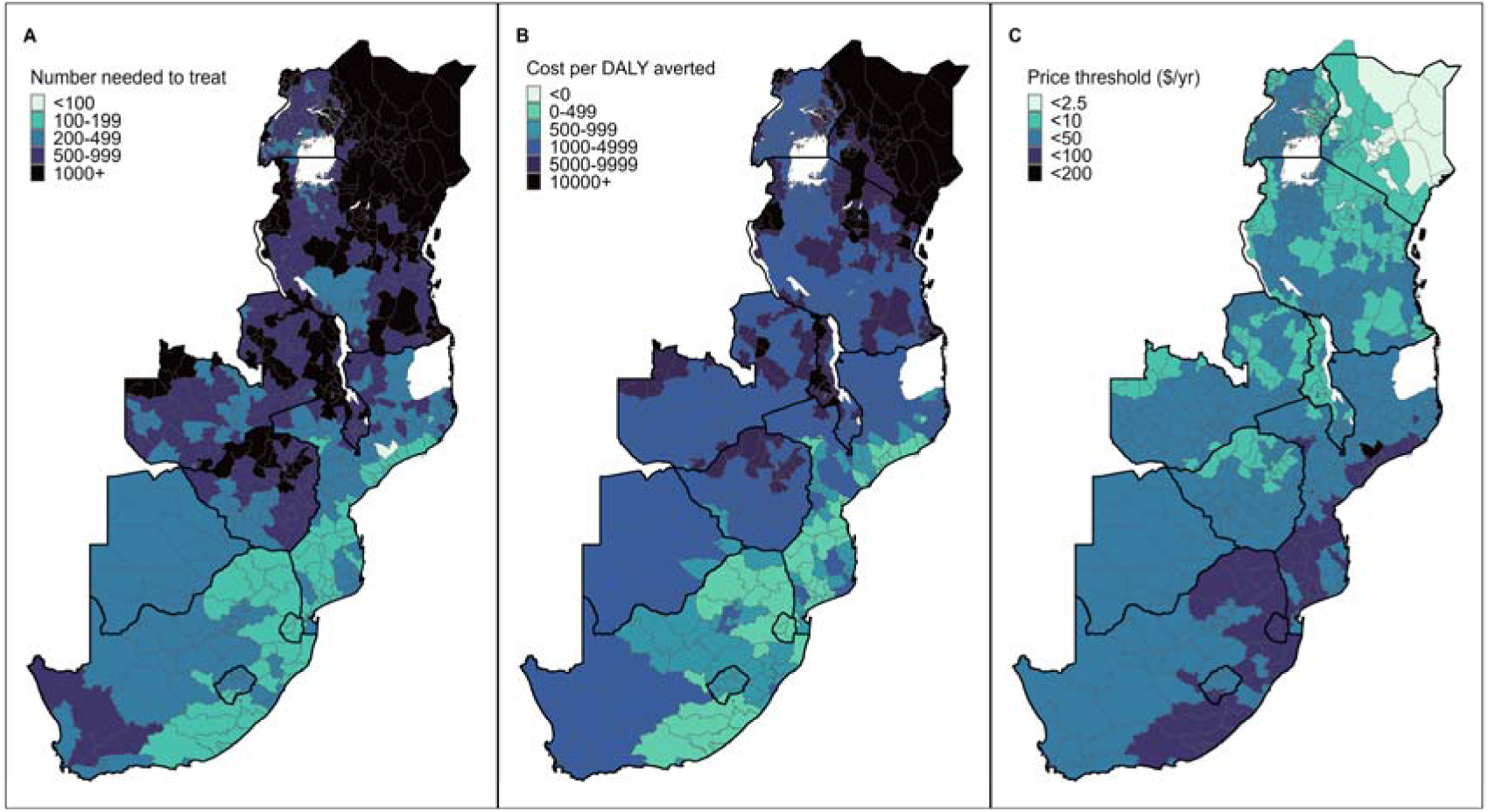
District-level costing metrics among adults aged 15–49. (A) Number needed to treat (person-years of PrEP to avert one infection), (B) Cost per DALY averted (at a fully-loaded Lenacapavir price of $55 pppy), (C) Price threshold to achieve cost effectiveness (at <$500 per DALY averted) among men and women 15–49 years of age by district.

To assess where uniform provision of Lenacapavir would be cost effective (our base-case assumption), we evaluated district-level cost-effectiveness offer to women aged 15–24 years across a range of Lenacapavir prices (Figure 3). At a price of $125 pppy, Lenacapavir offer to young women with average district-level risk was cost effective in only a small number of high-incidence districts: four districts in South Africa and eight in Mozambique (Figure 3A). At the lower price $55 pppy, the number of districts where provision was cost-effective expanded substantially, encompassing most districts in South Africa and Mozambique, along with select high-incidence districts in Botswana. Few districts in Tanzania, Uganda, Zambia, and Zimbabwe met cost-effectiveness at $55 pppy, and Kenya and Malawi had no districts that met cost effectiveness thresholds at that price. These patterns are summarized in Figure 3B, which shows the cumulative proportion of the population at risk residing in districts where provision was cost-effective as a function of Lenacapavir price. As prices decline, progressively lower-incidence districts are included, increasing population coverage but reducing average efficiency.

**Figure 3.**
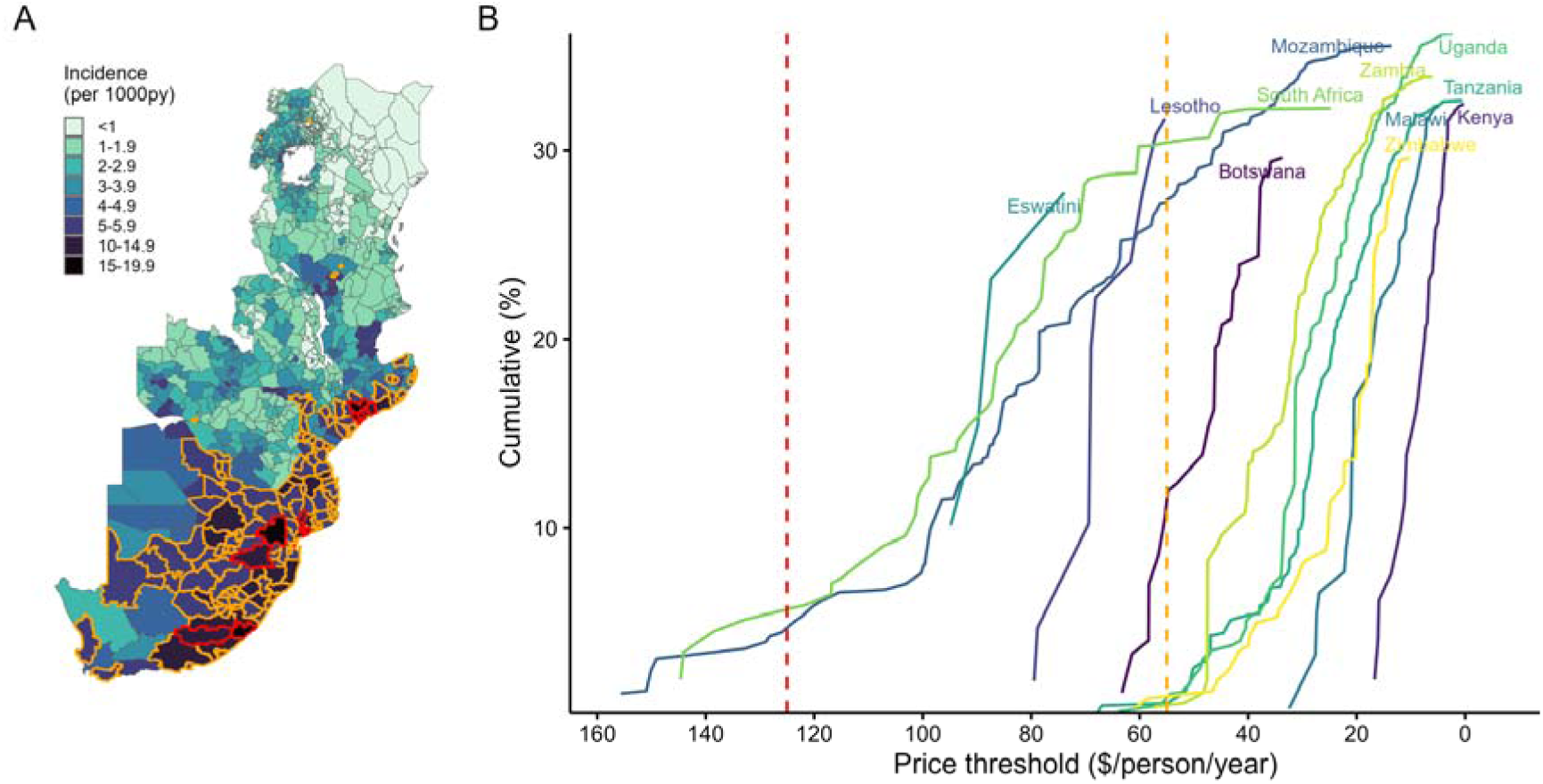
District-level incidence among women 15–24 years. Red outline represents districts in which universal offer of Len to young women is cost-effective ($/DALY averted <$500) at a Lenacapavir cost of $125 pppy versus $55 pppy (adding orange outlined districts). (B) Cumulative infections reached by price threshold for each country.

We focused on South Africa’s 52 districts to compare the impact and cost-effectiveness of district-level allocation strategies prioritizing different risk strata, defined as quartiles of a continuous underlying risk distribution (Table 2; Figure 4). When Lenacapavir provision was concentrated among individuals with incidence exceeding twice the district-level average, (consistent with the uptake assumptions for universal PrEP access), it achieved cost-effectiveness across a large number of districts. Among women aged 15–24 years in the highest-risk stratum (>2× district average incidence), mean incidence across districts was 21.5 per 1,000 person-years (range: 7.3–37.1). At $55 pppy, universally offer of Lenacapavir to this group would be cost-effective in all 52 districts, encompassing 4.1% of HIV-negative adults (aged 15–49 years) while reaching 18.2% of all new adult infections. This strategy achieved the lowest NNT across districts (mean 56; range 28–145) and the highest average ratio of infections averted per percentage of population covered (4.4). At $125 pppy, universal offer to women aged 15–24 with >2× risk remained cost-effective in 45 districts, encompassing 3.7% of HIV-negative adults and 17.1% of new infections. Patterns were comparable for other age and sex groups at lower prices: at $55 pppy, Lenacapavir was cost-effective in 47 of 52 districts among women aged 25–34 years, 47 districts among men aged 25–34 years, and 42 districts among women aged 35–49 years. In contrast, allocation to lower-risk subgroups (1–<2× district incidence) was rarely cost-effective at $125 pppy and only occasionally so at $55 pppy (Table 2).

**Figure 4.**
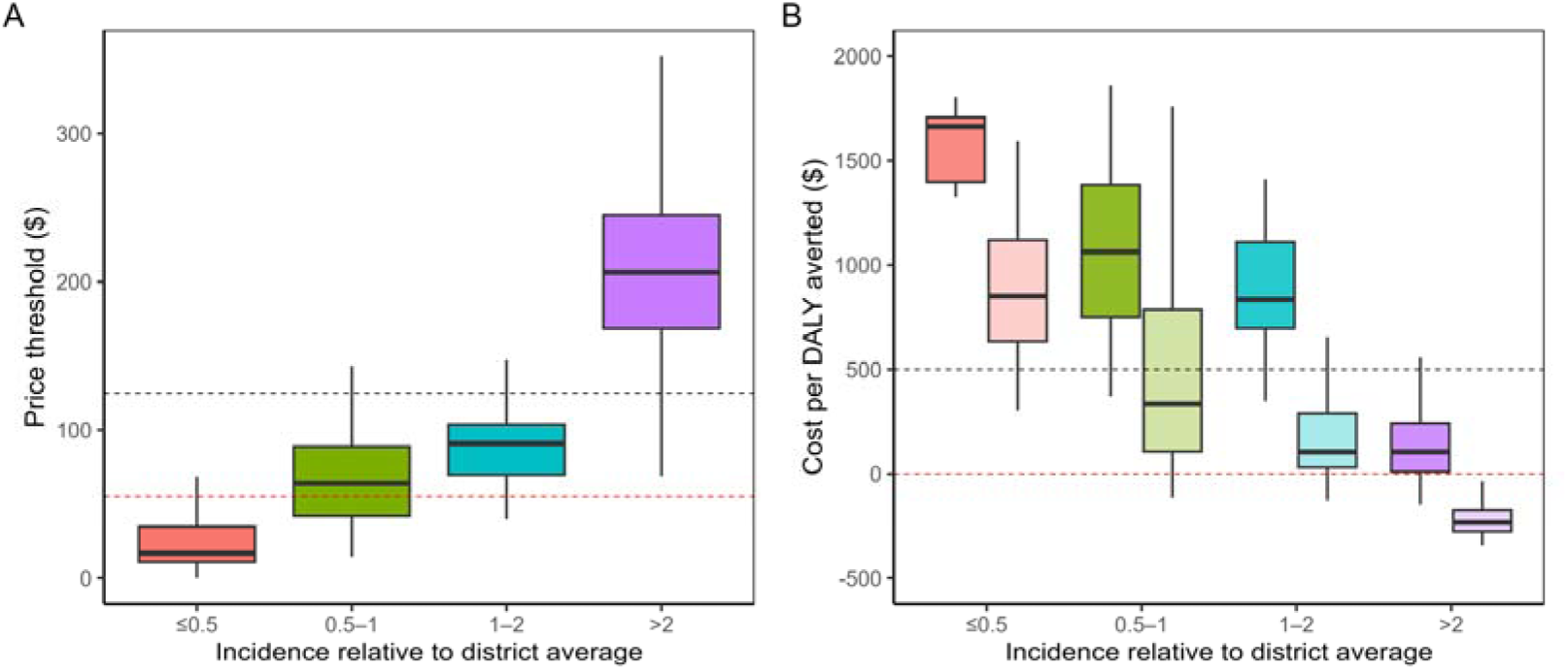
Sensitivity analysis showing how the relative difference in incidence between those who uptake Lenacapavir and the district average impacts the distribution (A) price thresholds and (B) costs per DALY averted at $125 pppy (dark shaded bars) and $55 pppy (light shaded bars) across all districts in South Africa among women 15–24. Black and red dotted lines in plot A indicate $125 and $55 price points, respectively, and in plot B indicate cost effectiveness threshold of <$500 per DALY averted and cost saving threshold of <$0 per DALY averted, respectively. Models were run under baseline DALYs per HIV infection (10). Panel B shows that if those who uptake Lenacapavir have >2 times the incidence as the district average (purple bars), it would be cost effective at $125 pppy in ∼75% of all districts and cost saving in ∼25%, and at $55 pppy it would be cost saving in all districts. See FigureS5 for additional sensitivity analysis with reduced DALYs per HIV infection (7.5).

**Table 2.**
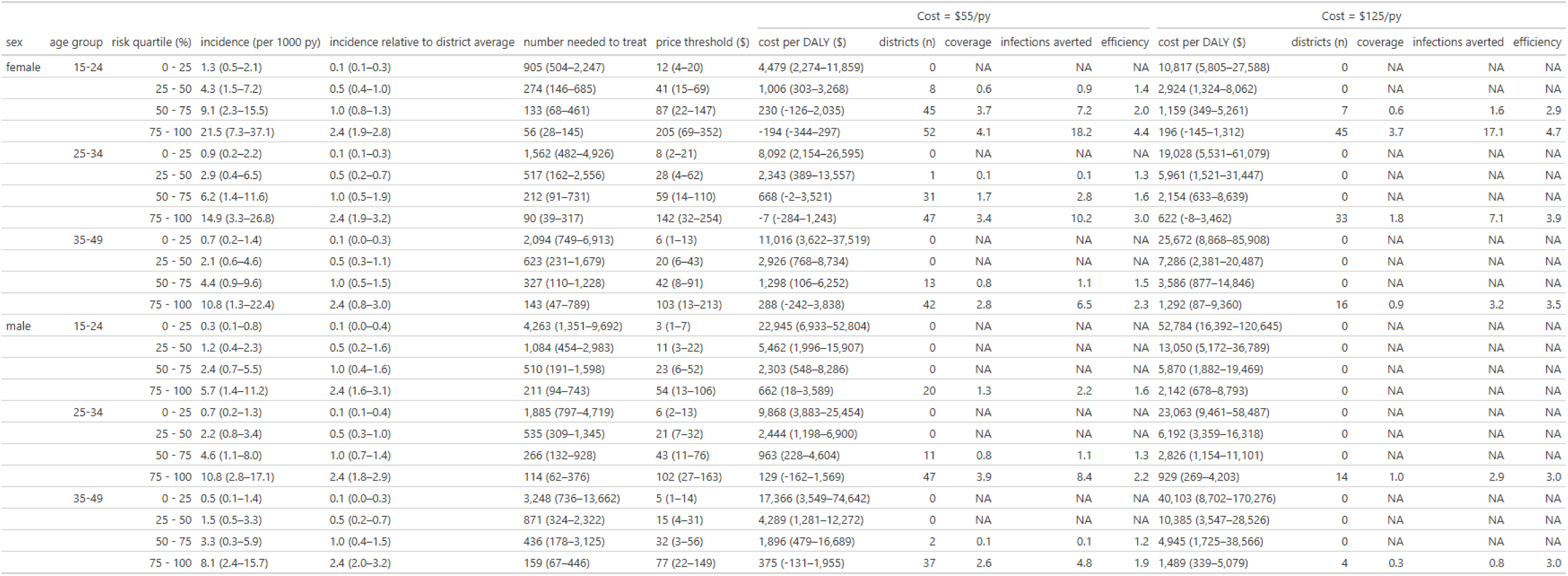
Model results comparing district-level Lenacapavir allocation strategies in 52 South African districts by age, sex, and risk-group prioritization (assuming base-case costing and DALY parameters). Under each costing scenario, “districts (n)” indicates the number of districts where the strategy is cost-effective ($/DALY averted < $500) and the resulting coverage and infections averted.

Figure 5 illustrates the cumulative impact of prioritizing Lenacapavir to South African districts with the highest age-, sex-specific incidence comparing average-risk versus higher-risk uptake scenarios. A higher-risk uptake assumption yielded large impact at low coverage: at 3% population coverage, approximately 15.5% of new infections could be reached, increasing to 23% at 5% coverage. In contrast, assuming average-risk uptake produced substantially lower impact at equivalent coverage levels. Achieving coverage of 25% of new infections required reaching 11% of the population under average-risk uptake, compared with only 5.5% coverage under higher-risk uptake. Estimates from the higher-risk uptake model were comparable to those from country-level dynamical transmission models prioritizing discrete high-risk groups and key populations in South Africa, where a 3–5% coverage yielded a 21–22% reduction in incidence^4,18^.

**Figure 5.**
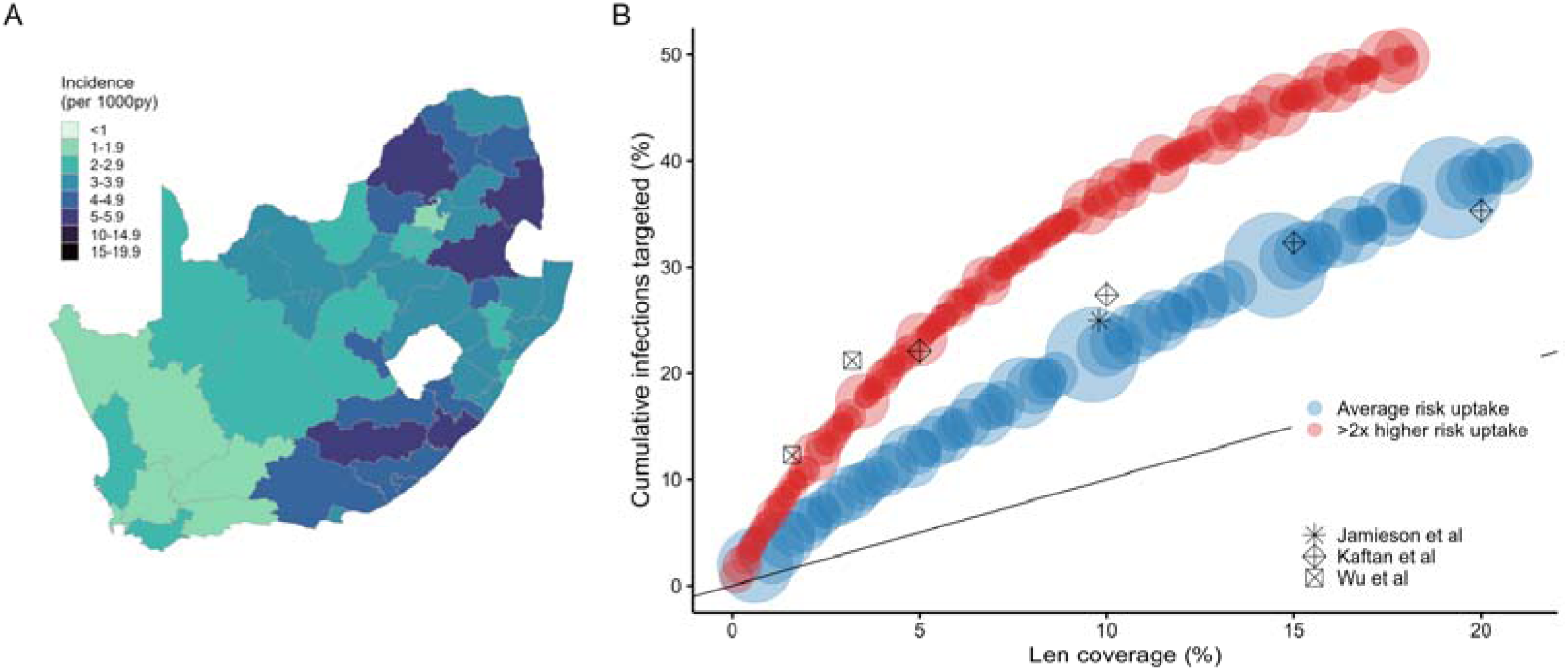
(A) Incidence (adults 15–49) map of South African districts (B) Cumulative percent of new infections within an increasingly inclusive HIV negative population (stratified by age and sex) comparing a scenario with average risk uptake versus uptake in those with >2x district-level risk. Symbols indicate coverage and impact estimates from prior mathematical modeling of Lenacapavir implementation strategies based on country level allocation prioritizing discrete high-risk populations (e.g., FSW, their clients, AGYW with 2+ sex partners, and men who have sex with men)^4,18,19^.

Figures 6a and 6b show, at fully loaded annual Lenacapavir costs of $125 and $55 pppy, respectively, visual representations of the South African districts in which a universal offer (assuming uptake in those with >2× district-level risk risk) is cost-effective, versus districts where further risk-based differentiation would be required to achieve cost-effective delivery. At $125 pppy, further risk prioritization would be needed to achieve cost effectiveness in most districts men of all age-groups and for women 35–49. For women 15–24 and 25–34, no further risk prioritization beyond universal offer would be required to achieve cost effectiveness in most districts at $125 pppy. The geographic scope where universal offer achieves cost-effectiveness expands considerably at $55 pppy. These patterns highlight substantial within-country heterogeneity and illustrate how district-level incidence thresholds can be used to distinguish settings where universal delivery is feasible from those where more risk-prioritized approaches are needed.

**Figure 6.**
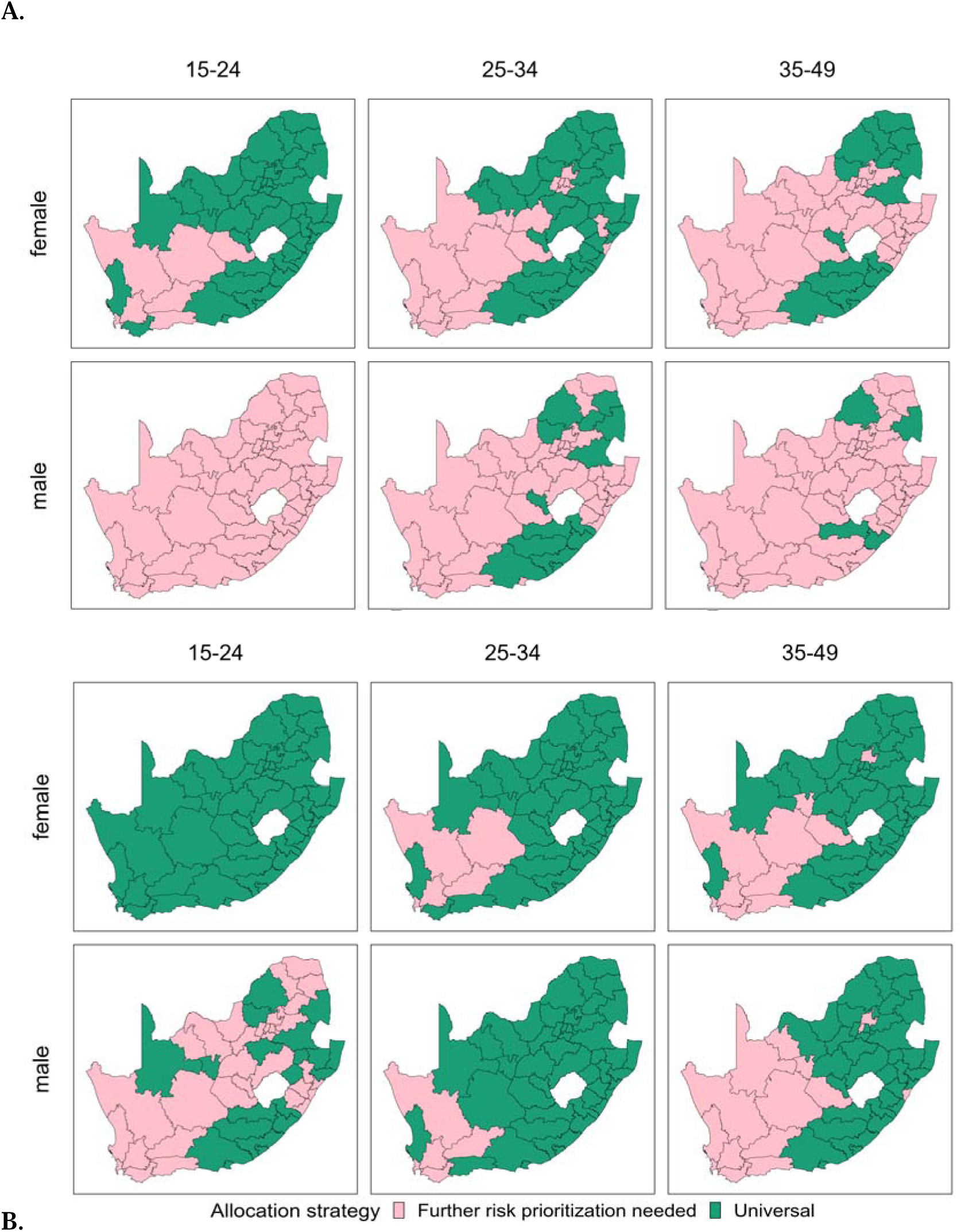
Districts in South Africa where a universal allocation strategy of Lenacapavir to women 15–24 years with assumed uptake among those with >2x district incidence would be cost-effective at < $500/DALY (green shaded areas) versus where delivery would require additional risk prioritization (pink shaded areas), comparing Lenacapavir costs of (A) $125 pppy and (B) $55 pppy. Results are stratified by age and gender

Sensitivity analyses showed that the spatial patterns of cost-effectiveness were robust to alternative DALY assumptions, with high-incidence districts consistently exhibiting lower costs per DALY averted and higher price thresholds for cost-effectiveness (Figures S3 and S4). While the number of districts meeting cost-effectiveness thresholds declined modestly at a more conservative assumption of 7.5 DALYs per HIV infection, the relative efficiency and prioritization ranking of age, sex, and risk groups remained unchanged. Lenacapavir remained cost-effective for universal offer to women aged 15–24 years in 42 districts at $125 pppy and in all 52 districts at $55 pppy (Table S1). At the lower DALY assumption, cost-effective delivery at $125 pppy was also observed for women aged 25–34 years (27 districts) and women aged 35–49 years (13 districts), with substantially broader cost-effectiveness across age and sex groups at $55 pppy. The risk strata parameter (uptake assumption) was still the largest determinant of impact and efficiency, with a sharp decline in both estimates when uptake is assumed in lower risk groups.

## Discussion

We applied a heuristic model based on district-level HIV incidence to project the direct impact and cost effectiveness geographic prioritization for Lenacapavir PrEP across districts in ESA. This simple model captures the substantial geographic and demographic heterogeneity in HIV incidence and can be used to compare the relative impact and cost-effectiveness of primary HIV prevention interventions across a wide range of settings and scenarios. Our findings highlight that universal delivery approaches are most efficient when implemented in geographically prioritized, high-incidence districts, particularly among younger women. In these settings, universal offer—under the assumption that those at highest risk will differentially uptake—achieves efficiency comparable to highly risk-based targeting strategies while avoiding the operational and social costs of behavioral screening. Universal allocation offers practical advantages for policymakers, who must make urgent decisions with constrained budgets and limited data.

Our projections of impact achieved through geographically targeted delivery with minimal risk stratification are broadly consistent with prior country-level analyses from South Africa using more complex dynamical models, highlighting the utility of a simple modeling approach. An agent-based transmission model from South Africa found that at $100 pppy, Lenacapavir would be cost effective if prioritized to high-risk groups and key populations, covering 3.2% of the population at risk resulting in a 21.2% reduction in incidence^4^. In another country-level, dynamical HIV model from South Africa, prioritizing Lenacapavir to those at highest risk (e.g, sex workers) and expanding allocation to lower-risk populations with increasing coverage levels (at 5, 10, 15, and 20%), resulted in corresponding reductions in incidence of 22.1, 27.4, 32.3, and 35.3%, respectively^18^. These estimates are consistent with our model’s estimated population-level effectiveness at reasonable Lenacapavir coverage levels (at 5% coverage our model estimates a >20% reduction in incidence). These findings suggest similar efficiency of a geographic approach to risk prioritization compared to a behavioral-risk approach.

Our analyses for South Africa highlighted the efficiency and cost-effectiveness of prioritizing uptake among sexually active young women 15–24 years, who represent the age group with highest incidence in South Africa. Other modelling analyses for South Africa have suggested greatest impact through prioritizing delivery to female sex workers, men who have sex with men, and pregnant and lactating women with higher risk^19^. These populations are not explicitly considered in our model, and therefore our results showing high impact and efficiency of prioritizing general population young women should not be interpreted as contradictory, but rather not directly comparable, to an approach that prioritizes discrete risk-groups and key populations. Implementation considerations may also affect attainable impact of different strategies; narrowly reaching key populations may be impactful, but impractical particularly in the context of recent dismantling of key population-focused services in South Africa^35^. On the other hand, pregnant women are reachable through ante-natal care (ANC) services but may have more limited prospects for self-selection for differential uptake among those with higher risk—which was key to our findings, and assumed in Jamieson *et al.’s* analysis^19^. Overall, all modelling analyses have tended to assume higher Lenacapavir uptake among those at higher risk in each population, but data quantifying this is lacking and a priority for upcoming implementation science studies to guide further strategic expansion.

A universal approach to Lenacapavir allocation guided by geographic indicators of risk overcomes several important limitations of delivery using risk screening tools based on highly personal sexual behavior questions: it reduces stigma, instills agency and trust in the individual, and may streamline PrEP implementation^12^. Real-world evidence supports the impact and efficiency of the universal offer approach^13–16^. However, as implementation expands into lower-incidence districts, efficiency diminishes, highlighting a trade-off between maximizing population coverage and maintaining cost-effectiveness. Our analyses were derived assuming HIV incidence rates as estimated for the year 2024. Reflecting the same intrinsic relationship between population incidence and cost-effectiveness, as HIV incidence continues to decline in future years, the set of districts where Lenacapavir provision is cost effective would be expected to shrink commensurately.

There are several limitations to our modeling approach that must be considered in the interpretation of our results. First, we assumed an average lifetime cost of HIV care ($5,000) and an average health loss of 10 DALYs associated with each HIV infection. DALYs per HIV infection are sensitive to assumptions about life expectancy, ART uptake and survival on treatment, and disability weights. Our estimates also exclude health losses and treatment costs avoided through secondary infections averted. While our cost-effectiveness results were sensitive to treatment costs and DALYs per HIV infection, universal offer of Lenacapavir remained cost effective in higher-incidence settings. As there is no expert consensus on cost-effectiveness threshold, we adopt a commonly cited threshold of $500 per DALY averted, which may not correspond with the explicit or implicit value used by decision makers in a specific country.

Secondly, our model estimates only the direct impact of Lenacapavir and does not include transmission dynamics needed to estimate secondary transmissions averted, including vertical transmissions averted by preventing infections in women of childbearing age. Therefore, our results are conservative. Further, our model does not explicitly consider key populations; instead, we assume a continuous distribution of risk within districts. While this may not fully capture the heterogeneity in discrete behavioral groups, it relies on fewer difficult-to-measure parameters and can complement more complex transmission models.

Together, these findings suggest that geographically prioritized and age- and sex-specific universal delivery of Lenacapavir can achieve substantial impact and cost-effectiveness in high-burden settings, even under conservative assumptions and in the absence of detailed individual-level risk data.

## Conclusion

Our analysis demonstrates that the impact and cost-effectiveness of Lenacapavir vary substantially across geographic settings, age groups, and risk strata, with the greatest efficiency achieved in high-incidence districts and among younger women. Mapping these patterns provides a practical framework for identifying where universal provision to specific demographic groups can be implemented cost-effectively and where additional prioritization may be required. Potential for lower Lenacapavir commodity and delivery costs expands the set of districts in which universal delivery is cost-effective, enabling decision-makers to balance efficiency and coverage under constrained budgets. These results offer actionable guidance for the phased and geographically targeted implementation of Lenacapavir in eastern and southern Africa.

## Data Availability

All data produced are available online at https://naomi-spectrum.unaids.org. Scripts used to generate datasets, run country simulations and models, and reproduce all tables and figures in the manuscript can be found at https://github.com/aakullian/LenOptim/tree/master/R/Manuscript

https://naomi-spectrum.unaids.org

## Supplementary materials

**Table S1.** Sensitivity analysis (reduced DALYs per HIV infection to 7.5) with model results comparing district-level Lenacapavir allocation strategies across 52 districts in South Africa by age, sex, and risk-group prioritization.

| sex | age group | risk quartile (%) | incidence (per 1000 py) | incidence relative to district average | number needed to treat | price threshold (\$) | Cost = \$55/py | | | | | Cost = \$125/py | | | | |
| --- | --- | --- | --- | --- | --- | --- | --- | --- | --- | --- | --- | --- | --- | --- | --- | --- |
| | | | | | | | cost per DALY (\$) | districts (n) | coverage | infections averted | efficiency | cost per DALY (\$) | districts (n) | coverage | infections averted | efficiency |
| female | 15-24 | 0 - 25 | 1.3 (0.5–2.1) | 0.1 (0.1–0.3) | 905 (504–2,247) | 11 (4–17) | 5,973 (3,033–15,811) | 0 | NA | NA | NA | 14,423 (7,741–36,784) | 0 | NA | NA | NA |
|  |  | 25 - 50 | 4.3 (1.5–7.2) | 0.5 (0.4–1.0) | 274 (146–685) | 36 (13–60) | 1,342 (404–4,357) | 4 | 0.4 | 0.6 | 1.4 | 3,898 (1,766–10,750) | 0 | NA | NA | NA |
|  |  | 50 - 75 | 9.1 (2.3–15.5) | 1.0 (0.8–1.3) | 133 (68–461) | 76 (19–129) | 307 (–169–2,713) | 44 | 3.6 | 7.2 | 2.0 | 1,545 (466–7,015) | 2 | 0.3 | 0.8 | 3.1 |
|  |  | 75 - 100 | 21.5 (7.3–37.1) | 2.4 (1.9–2.8) | 56 (28–145) | 179 (60–308) | –258 (–458–396) | 52 | 4.1 | 18.2 | 4.4 | 261 (–193–1,749) | 42 | 3.4 | 16.2 | 4.8 |
|  | 25-34 | 0 - 25 | 0.9 (0.2–2.2) | 0.1 (0.1–0.3) | 1,562 (482–4,926) | 7 (2–18) | 10,790 (2,872–35,460) | 0 | NA | NA | NA | 25,371 (7,375–81,439) | 0 | NA | NA | NA |
|  |  | 25 - 50 | 2.9 (0.4–6.5) | 0.5 (0.2–0.7) | 517 (162–2,556) | 24 (3–54) | 3,124 (519–18,076) | 0 | NA | NA | NA | 7,949 (2,028–41,930) | 0 | NA | NA | NA |
|  |  | 50 - 75 | 6.2 (1.4–11.6) | 1.0 (0.5–1.9) | 212 (91–731) | 52 (12–97) | 891 (–2–4,695) | 23 | 1.3 | 2.3 | 1.8 | 2,873 (844–11,519) | 0 | NA | NA | NA |
|  |  | 75 - 100 | 14.9 (3.3–26.8) | 2.4 (1.9–3.2) | 90 (39–317) | 124 (28–222) | –9 (–378–1,658) | 46 | 3.4 | 10.1 | 3.0 | 829 (–11–4,616) | 27 | 1.6 | 6.3 | 4.1 |
|  | 35-49 | 0 - 25 | 0.7 (0.2–1.4) | 0.1 (0.0–0.3) | 2,094 (749–6,913) | 6 (1–12) | 14,688 (4,829–50,026) | 0 | NA | NA | NA | 34,230 (11,824–114,544) | 0 | NA | NA | NA |
|  |  | 25 - 50 | 2.1 (0.6–4.6) | 0.5 (0.3–1.1) | 623 (231–1,679) | 18 (5–38) | 3,901 (1,024–11,646) | 0 | NA | NA | NA | 9,715 (3,175–27,316) | 0 | NA | NA | NA |
|  |  | 50 - 75 | 4.4 (0.9–9.6) | 1.0 (0.5–1.5) | 327 (110–1,228) | 36 (7–79) | 1,731 (141–8,337) | 10 | 0.6 | 1.0 | 1.6 | 4,782 (1,170–19,795) | 0 | NA | NA | NA |
|  |  | 75 - 100 | 10.8 (1.3–22.4) | 2.4 (0.8–3.0) | 143 (47–789) | 90 (11–186) | 385 (–322–5,118) | 41 | 2.7 | 6.5 | 2.4 | 1,722 (117–12,480) | 13 | 0.8 | 2.7 | 3.7 |
| male | 15-24 | 0 - 25 | 0.3 (0.1–0.8) | 0.1 (0.0–0.4) | 4,263 (1,351–9,692) | 3 (1–6) | 30,593 (9,243–70,405) | 0 | NA | NA | NA | 70,379 (21,856–160,860) | 0 | NA | NA | NA |
|  |  | 25 - 50 | 1.2 (0.4–2.3) | 0.5 (0.2–1.6) | 1,084 (454–2,983) | 10 (3–19) | 7,283 (2,661–21,210) | 0 | NA | NA | NA | 17,400 (6,896–49,053) | 0 | NA | NA | NA |
|  |  | 50 - 75 | 2.4 (0.7–5.5) | 1.0 (0.4–1.6) | 510 (191–1,598) | 20 (5–46) | 3,070 (731–11,049) | 0 | NA | NA | NA | 7,827 (2,509–25,959) | 0 | NA | NA | NA |
|  |  | 75 - 100 | 5.7 (1.4–11.2) | 2.4 (1.6–3.1) | 211 (94–743) | 47 (12–93) | 883 (24–4,785) | 16 | 1.2 | 1.9 | 1.7 | 2,856 (904–11,724) | 0 | NA | NA | NA |
|  | 25-34 | 0 - 25 | 0.7 (0.2–1.3) | 0.1 (0.1–0.4) | 1,885 (797–4,719) | 6 (2–11) | 13,157 (5,177–33,939) | 0 | NA | NA | NA | 30,751 (12,615–77,982) | 0 | NA | NA | NA |
|  |  | 25 - 50 | 2.2 (0.8–3.4) | 0.5 (0.3–1.0) | 535 (309–1,345) | 19 (7–28) | 3,259 (1,597–9,200) | 0 | NA | NA | NA | 8,255 (4,478–21,757) | 0 | NA | NA | NA |
|  |  | 50 - 75 | 4.6 (1.1–8.0) | 1.0 (0.7–1.4) | 266 (132–928) | 38 (9–66) | 1,284 (304–6,139) | 3 | 0.2 | 0.3 | 1.5 | 3,767 (1,539–14,801) | 0 | NA | NA | NA |
|  |  | 75 - 100 | 10.8 (2.8–17.1) | 2.4 (1.8–2.9) | 114 (62–376) | 90 (23–142) | 172 (–216–2,092) | 45 | 3.5 | 7.9 | 2.3 | 1,239 (358–5,604) | 6 | 0.5 | 1.5 | 3.2 |
|  | 35-49 | 0 - 25 | 0.5 (0.1–1.4) | 0.1 (0.0–0.3) | 3,248 (736–13,662) | 4 (1–12) | 23,154 (4,732–99,522) | 0 | NA | NA | NA | 53,471 (11,602–227,035) | 0 | NA | NA | NA |
|  |  | 25 - 50 | 1.5 (0.5–3.3) | 0.5 (0.2–0.7) | 871 (324–2,322) | 13 (4–27) | 5,719 (1,707–16,362) | 0 | NA | NA | NA | 13,846 (4,729–38,035) | 0 | NA | NA | NA |
|  |  | 50 - 75 | 3.3 (0.3–5.9) | 1.0 (0.4–1.5) | 436 (178–3,125) | 28 (3–49) | 2,528 (639–22,252) | 0 | NA | NA | NA | 6,593 (2,300–51,421) | 0 | NA | NA | NA |
|  |  | 75 - 100 | 8.1 (2.4–15.7) | 2.4 (2.0–3.2) | 159 (67–446) | 67 (20–130) | 500 (–174–2,606) | 32 | 1.9 | 4.0 | 2.1 | 1,985 (453–6,772) | 1 | 0.0 | 0.1 | 3.2 |

**Figure S1.**
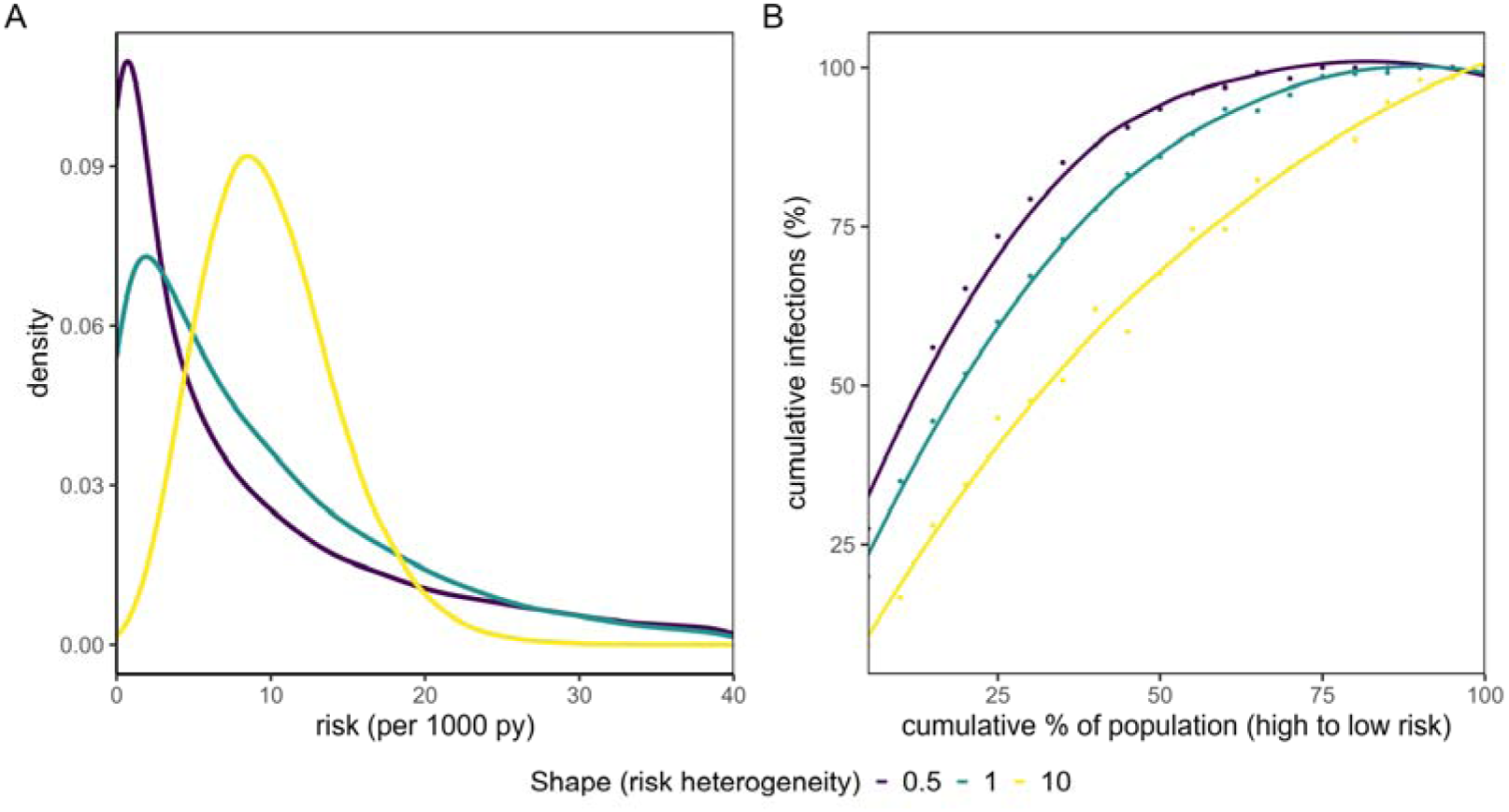
(A) Continuous risk distribution from a Gamma function, varying the shape parameter (0.1, 1, 10), with mean fixed at an incidence of 10/1000py, (B) Cumulative infections averted by increasingly inclusive coverage of the population, from highest to lowest risk)

**Figure S2.**
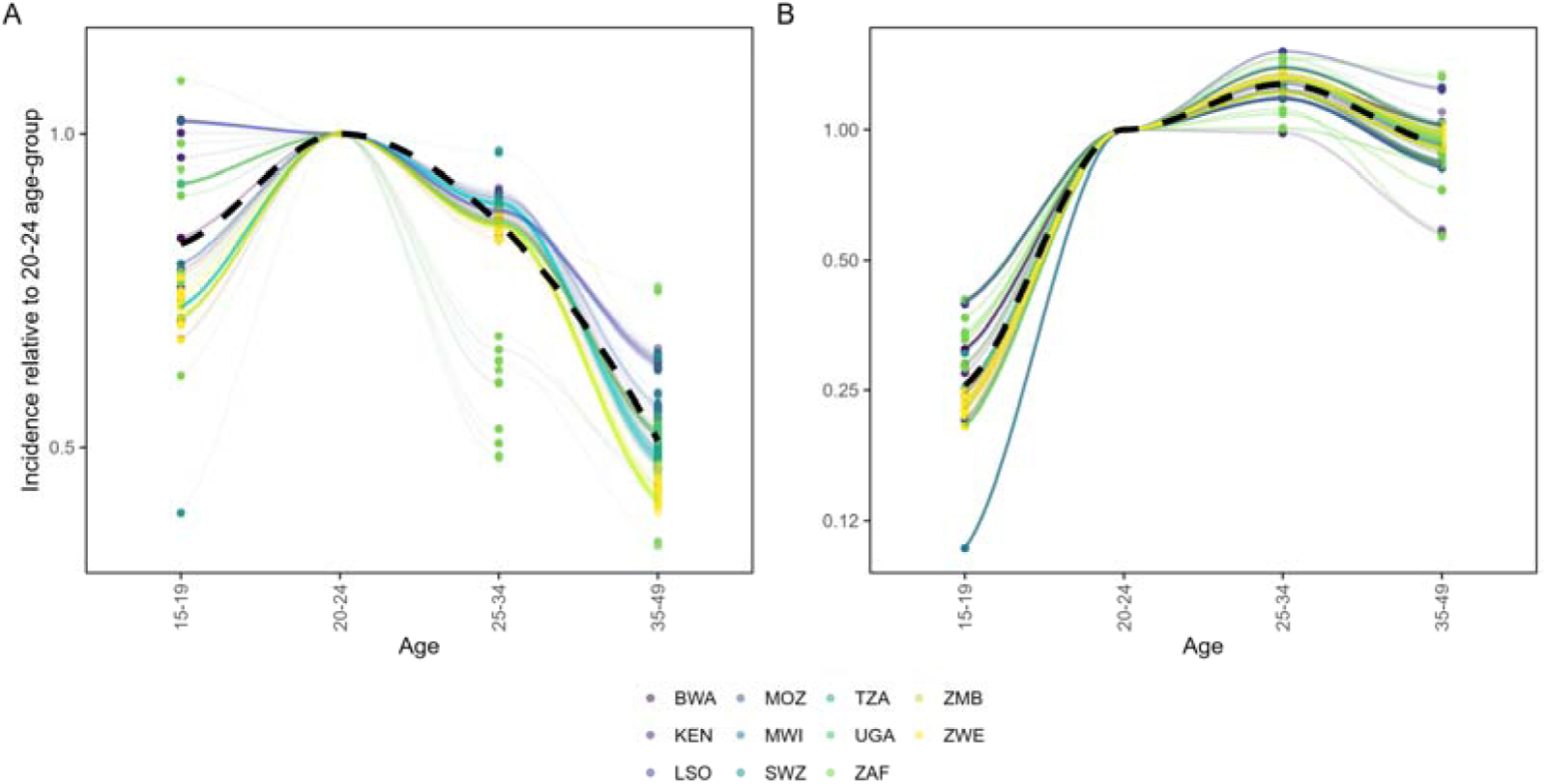
District-level HIV incidence by age relative to 20–24 y/o’s among: (A) Women and (B) Men. Loess smooth across all districts indicated with a black dashed curve. Note y-axis is on a log2 scale and axis extents are different

**Figure S3.**
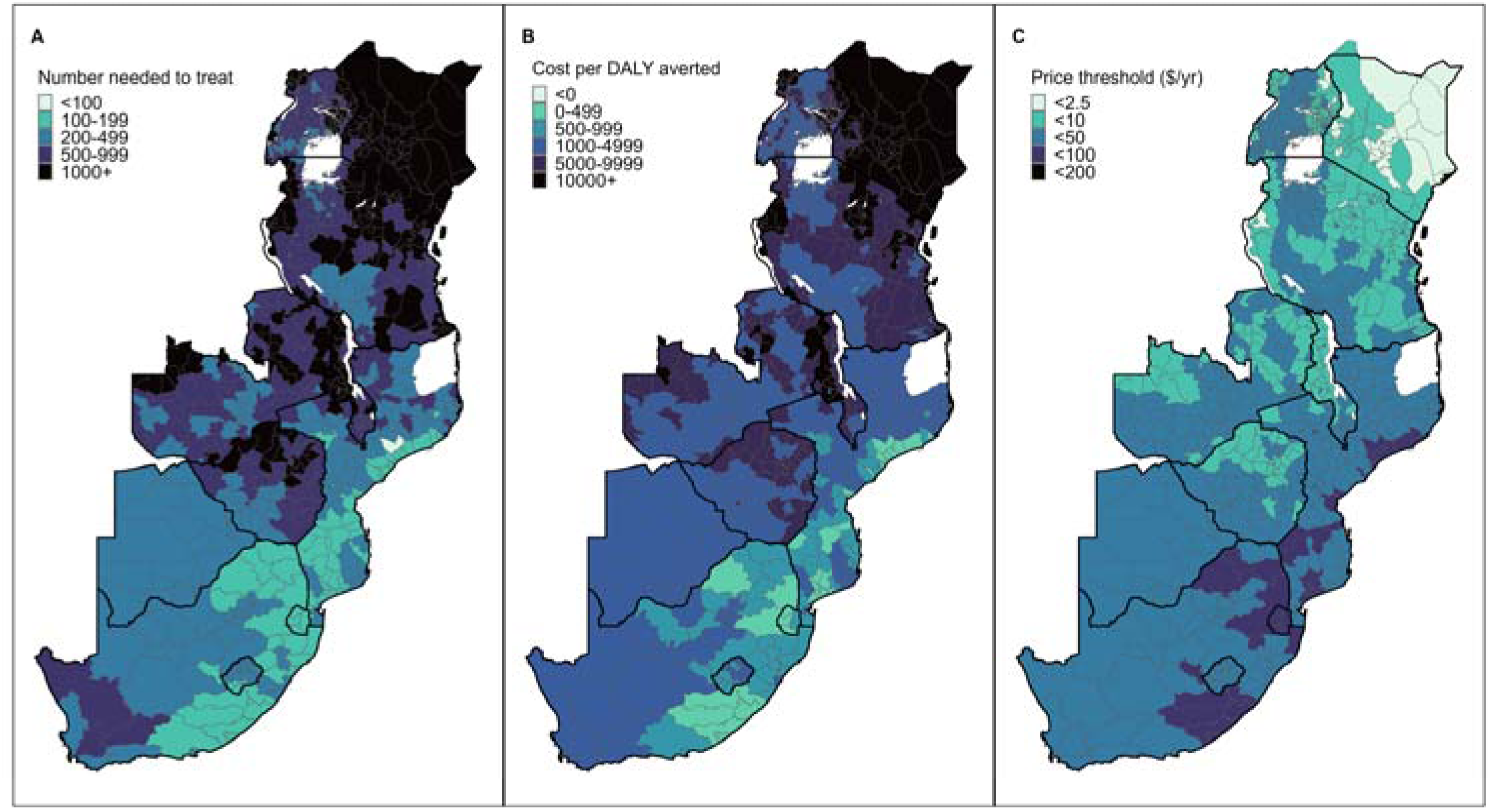
Sensitivity plot with models run under lower estimates of 7.5 DALYs per HIV infection. (A) Number needed to treat (person-years of PrEP to avert one infection), (B) Cost per DALY averted, (C) Price threshold to achieve cost effectiveness (at $500 per DALY averted) among women 15–24 years of age by district.

**Figure S4.**
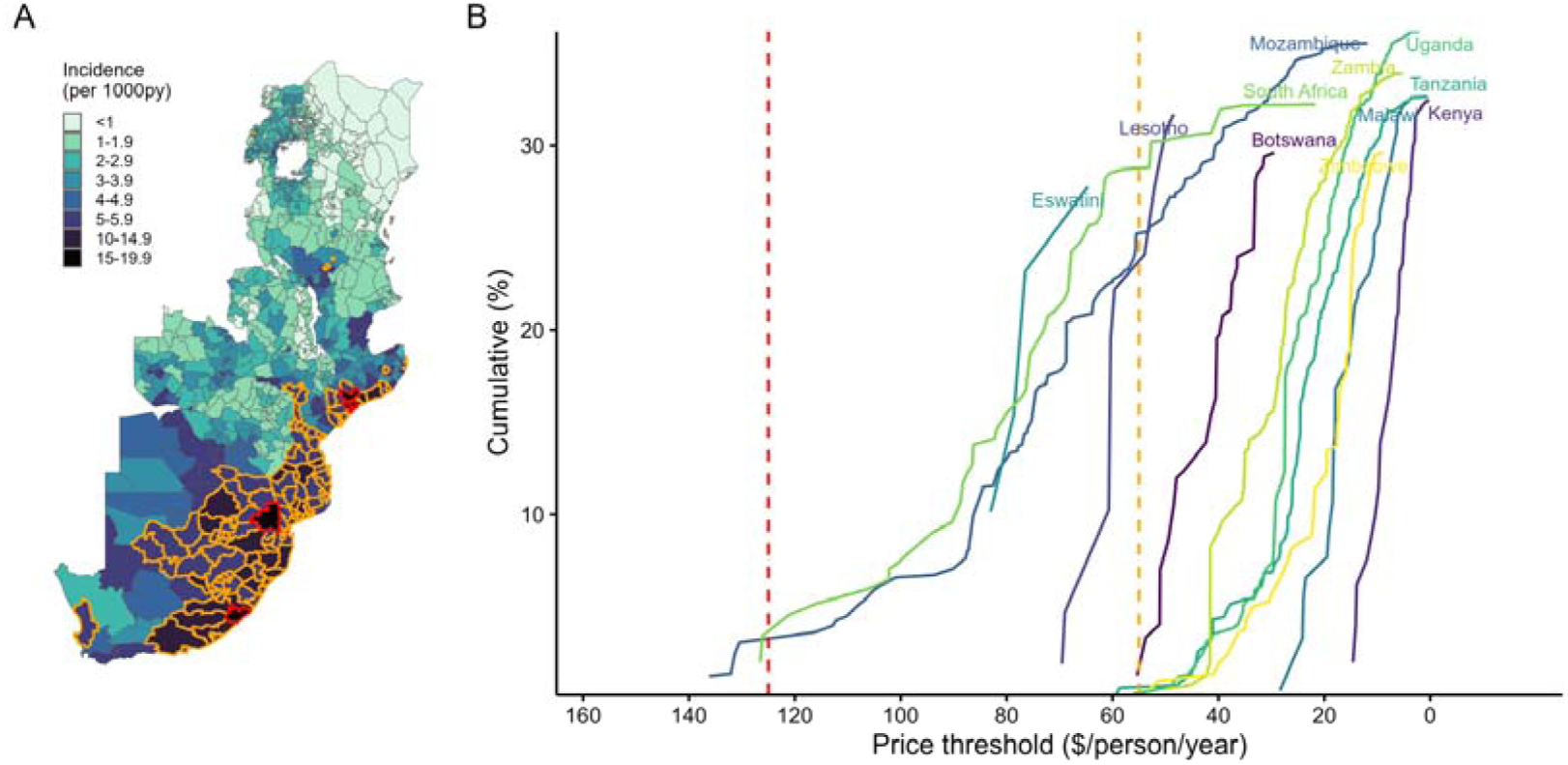
Sensitivity plot with models run under lower estimates of 7.5 DALYs per HIV infection. (A) District-level incidence among women 15–24 with cost-effective districts outlined in red (at Len cost of $125 pppy) and red + orange (at Len cost of $55 pppy). (B) Cumulative infections averted with increasingly inclusive set of districts (from high to low incidence) by price threshold for each country. Red dashed line indicates $125 pppy Lenacapavir cost and orange dashed line indicates $55 pppy.

**Figure S5.**
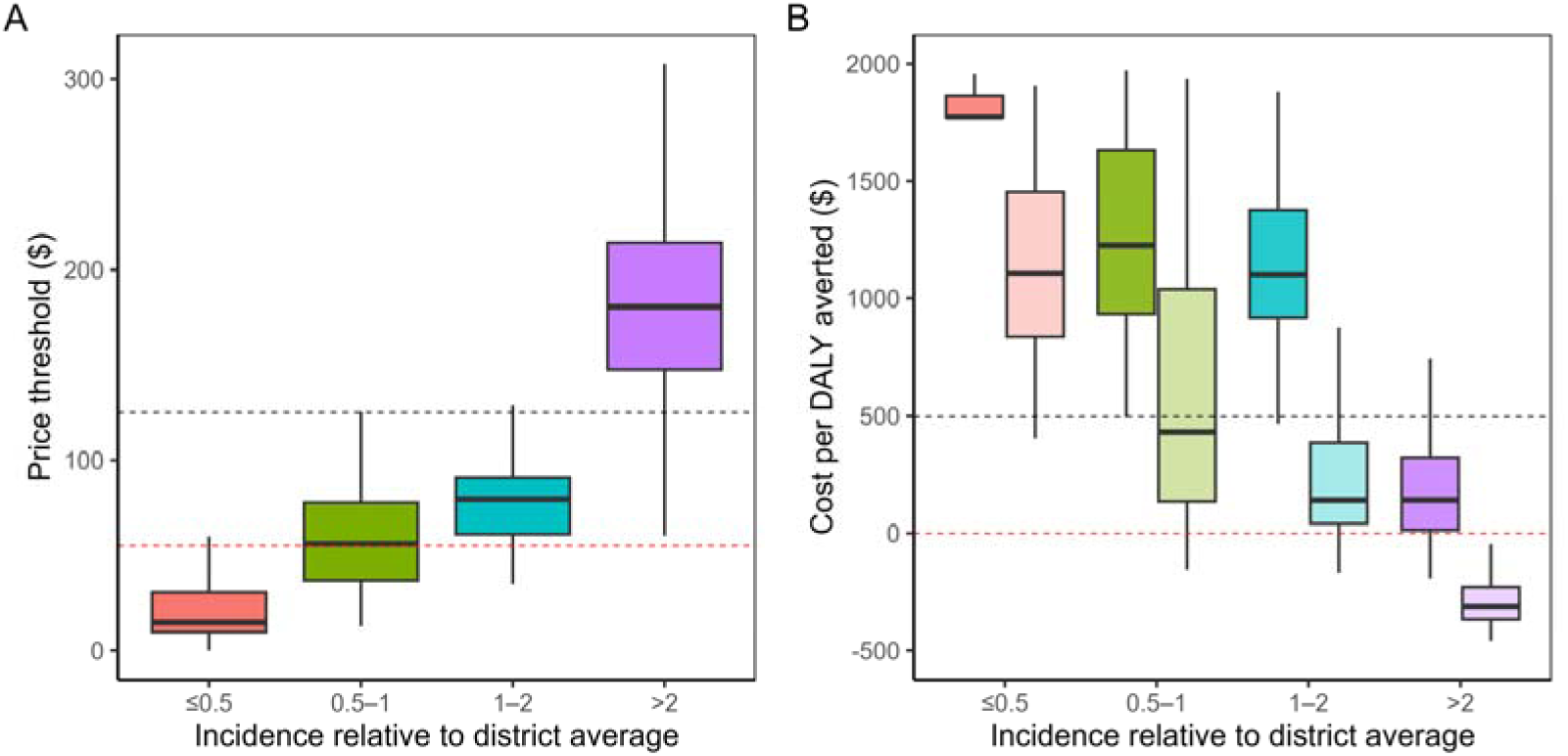
Sensitivity analysis plots under reduced DALYs per HIV infection (7.5) with (A) Distribution of price thresholds (black dashed line indicate $125 pppy and red line indicates $55 pppy) and (B) costs per DALY averted at $125 pppy (dark shaded bars) and $55 pppy (light shaded bars) for all districts in South Africa among women 15–24 by relative difference in incidence between those who uptake Lenacapavir and the district average. Black and red dotted lines in plot A indicate $125 and $55 < $500 per DALY averted and cost saving threshold of <$0 per DALY averted, respectively.

The following equations are used in this analysis:

**1. Number needed to treat (NNT):**

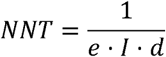

Where:

- *e*: efficacy of the intervention
- *I*: HIV incidence in the absence of intervention
- *d*: duration of protection (in years)

**2. Cost-effectiveness (cost per DALY averted):**

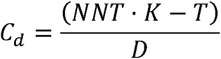

Where:

- *K*: cost per unit of prevention
- *T*: lifetime treatment cost of HIV
- *D*: DALYs associated with HIV infection

**3. Maximum price per dose (price threshold):**

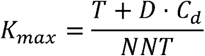

**4. Annual infections averted (impact):**

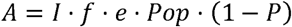

Where:

- *f*: coverage of target population
- *P*: HIV prevalence
- *Pop*: total population

All calculations were stratified by age, sex, geography, and assumed different prioritization scenarios.

